# Universal scaling law for COVID-19 propagation in urban centers

**DOI:** 10.1101/2020.06.22.20137604

**Authors:** Ben-Hur Francisco Cardoso, Sebastián Gonçalves

## Abstract

Due to the COVID-19 pandemic, there is a high demand for Susceptible-Infective-Recovered (SIR) models to adjust and predict the number of cases in urban areas. Forecasting, however, is a difficult task, because the change in people’s behavior reflects in a continuous change in the parameters of the model. An important question is what we can use from one city to another; if what happened in Madrid could have been applied to New York and then, if what we have learned from this city would be useful for São Paulo. To answer this question, we present an analysis of the transmission rate of COVID-19 as a function of population density and population size for US counties, cities of Brazil, German, and Portugal. Contrary to the common hypothesis in epidemics modeling, we observe a higher disease transmissibility for higher city’s population density/size –with the latter showing more predicting power. We present a contact rate scaling theory that explain the results, predicting that the basic reproductive number *R*_0_ of epidemics scales as the logarithm of the city size.

## Introduction

The epidemic of COVID-19 that started in the Chinese city of Wuhan in December of 2019, was declared a pandemic on March 11th, 2020 by the World Health Organization (WHO). While in many European countries (1) the epidemic is slowing down after months of restrictions, the US is still struggling with the highest numbers of active cases and deaths. Presently, however, the epicenter is in Brazil, where the epidemic has the potential to hit even worse than in the US. The COVID-19 is a human contact driven disease, therefore it is necessary to work at the municipal level (2, 3) to get precise predictions. Such an approach, in addition to providing more accurate forecasts, it would allow us to apply the experience of one city to others. With this idea in mind, we focus on the *basic reproductive number R*_0_, a key concept in the mathematical description of epidemics, that measure the average number of secondary cases generated by each infected subject.

For a better understating, we factorize the basic reproductive number as *R*_0_ = *τpC*, where *τ* is the infectious period, *C* is the *per capita* contact rate and *p* the transmission probability. The probability of infection *p* ^1^ *and the infectious period τ* are characteristic of the disease itself. Therefore, to make the comparison between cities, it is necessary to understand how the contact rate is related to demographics.

In epidemiological modeling, the *mass action* hypothesis (4) is generally followed: the *per capita* contact rate is assumed invariant of city size. Our analysis shows, however, that this assumption cannot explain the available COVID-19 data. Alternatively, we propose a scaling theory that explains the dependence between contact rate and urban size or density, which then explain how the epidemic growth rate (*i*.*e. R*_0_) depends on size or density.

### Population size and Population density

Consider a city with population size *N* and land area *A*. There are two main competing hypothesis that try to explain how the *per capita* contact rate *C* varies with *N* and *A*: the population **size driven** contact rate, where *C* = *C*(*N*); and the population **density driven** contact rate, stating that *C* = *C*(*ρ*), where *ρ* = *N/A*. While the first approach assumes that the social mobility network grows in larger areas, allowing more distant people to interact (5), the second one assumes that the length traveled by the individuals is invariant of the size of the city (6).

Intriguingly, based on data of disease transmission in the United States, both approaches appear to be valid (6–8). The reason for this is the quasi-linear relation between the density and size population of the US counties, as shown in Fig. 1. Indeed, we have found that the best fit is *ρ*∝*N* ^*λ*^, with *λ*≈1.03. Assuming a linear relation instead, *ρ* = *kN* gives an equally valid fit, with *k* = 0.00059 Km^−2^ (this gives a typical US county diameter of 46.5 Km). An almost constant density across counties, or no correlation between *ρ* and *N*, cannot explain the data.

**Fig. 1.**
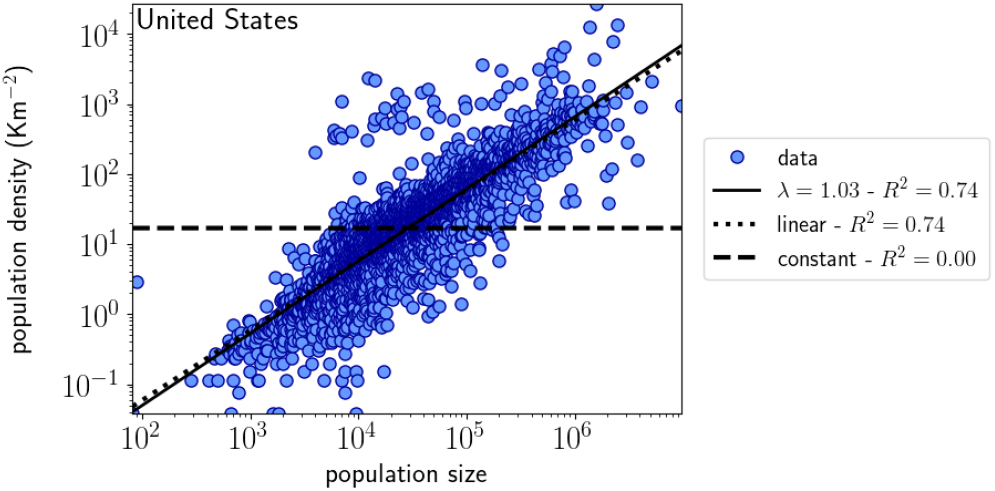
Population density (*ρ*) *vs* population size (*N*) for US counties, represented by blue dots (data). The solid line is a fit to a power law, *ρ* ∝*N* ^*λ*^, while the dotted line is a fit to the *ρ* = *kN* relation, with *k* = 0.00059 Km^−2^. Both fits have equal correlation coefficient, *R* = 0.86. The dashed line represents the average density. See Materials and Methods section for data source and fit methodology.

In addition to the US, we study the COVID-19 transmission in cities of Portugal, Brazil and Germany. While the same quasi-linear scaling *λ* = 1.03 was found for Portugal’s municipalities, in the other two countries the population density of cities does not correlate well with their size. The linear fit is strong for Portugal, weak for Brazil and almost nonexistent for Germany. Constant density also cannot explain the data for these countries (see Figs. S1, S3 and S2 in Supplementary Information Appendix).

Since the urban size and density are not well correlated in Brazil and Germany, we use these countries to verify which of the approaches, size-driven or density-driven, is the valid one. The result may be useful during the present COVID-19 pandemic and for futures ones.

### Contact rate scaling theory

Consider *N ≫* 1 individuals –represented as nodes– distributed uniformly in a two-dimensional space with land area A. As introduced by Noulas *et al* (9), we can expect an individual *i* to form a link with individual *j* with probability

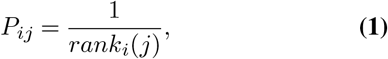

where *rank*_*i*_(*j*) is the number of neighbors closer to *i* than *j*. Assuming that the distance between these two individuals is *r*, we then have

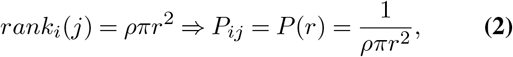

with *ρ* = *N/A* as the population density. First, since 0 ≤ *P* ≤1, we must impose a bottom cutoff radius *r*_0_ such that

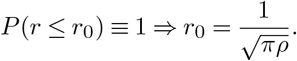

Secondly, it is natural to assume an upper cutoff radius *r*_1_ for long distances such that *P* (*r > r*_1_) = 0. Then, similarly to *Pan et al* (6), the mean degree of the social interaction network is

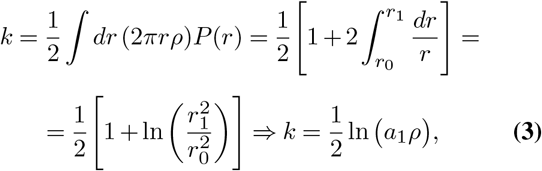

where 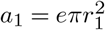 is the coverage area of individual mobility and the 1/2 factor eliminates the double counting.

It is well reasonable to expect the *per capita* contact rate *C*, and so *R*_0_, to be proportional to the mean degree *k*. Therefore, following the population density driven approach, which states that *a*_1_ is invariant, from Eq. 3 we arrive at

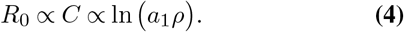

On the other hand, with the population size-driven approach, the mobility increases with *A* (*a*_1_ = *κA*), then from Eq. 3 we now have

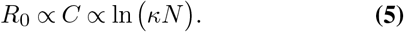

## Results

In this Section we check on the validity of the scaling model (Eqs. 4 and 5) in comparison to the traditional *mass action* hypothesis. In the Materials and Methods section, we explain how we estimate the basic reproductive number *R*_0_ of 1931 counties of the United States, 546 cities of Brazil, 401 cities of Germany and, 143 municipalities of Portugal, based on the COVID-19 related data. Finally, we give a plausible explanation for the marginal deviation of data from the scaling based on available data of mobility.

### Population density driven contact rate

In Fig. 2 we show the relation between *R*_0_ and the population density for different counties of the United States. We see that the proposed model (density driven, Eq. 4) provides a good fit, while the null (mass action) hypothesis cannot explain the data. A similar behaviour was found for Portugal (see Fig. S4 in Supplementary Information Appendix).

**Fig. 2.**
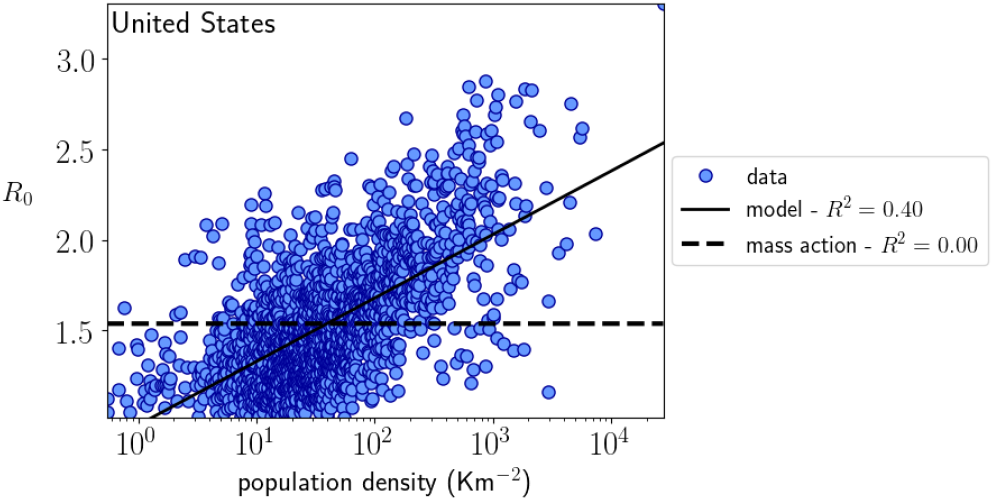
*R*_0_ *vs* the population density for different counties of the United States (see Materials and Methods section for fit methodology).

However, this behavior seems not to be universal. In the cases of Brazil and Germany the density driven model has almost the same predictability that the mass action hypothesis (see Figs. S5 and S6 in Supplementary Information Appendix). Yet, the *per capita* contact rate may scale with the population size instead of the population density as we see in the next Subsection.

### Population size driven contact rate

In Fig. 3, we show the relation between *R*_0_ and population size for urban regions of the United States, as well as for Brazil, Germany, and Portugal. We can see that the model (Eq. 5) is the one that provides a reasonable universal fit, while again the mass action hypothesis fails to explain the results (see Figs. S7, S9 and S10 in Supplementary Information Appendix for country-separated plots).

**Fig. 3.**
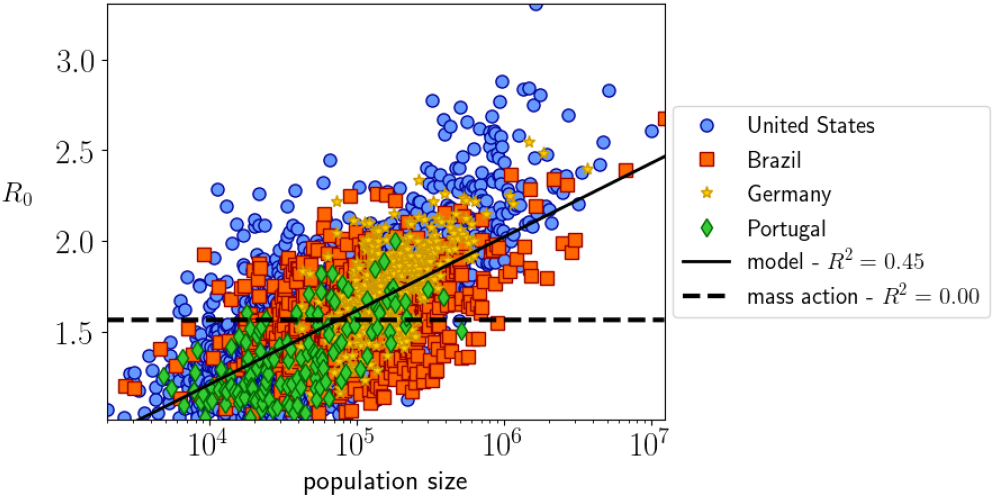
*R*_0_ *vs* the population size for urban regions of four different countries: the US, Brazil, Germany, and Portugal (see Materials and Methods section for fit methodology).

How can we explain that the COVID-19 data for the US and Portugal can be described by either one of the two hypothesis, the density driven and the size driven? This apparent ambiguity relies in the linear relation between population size and population density (*ρ* ∝ *N*) observed in the urban centers for these two countries (Section *Population size and Population density*). However, for Brazil and Germany centers, only the size driven approach appears to explain the scaling; thus, we conclude that the *size* driven approach has a great explanatory power than the *density* driven hypothesis.

### Marginal deviation and mobility reduction

The dispersion of points above or below the average – given by Eq. 5 – corresponds to areas where the epidemic spreads faster or slower than the average. We see in Fig. 3 that for the same population size we have municipalities with higher or lower spread, which besides the different procedures in reporting data, it could be telling something about the differences in dealing with the epidemic in different places.

Thus, we compare the marginal deviation of *R*_0_ (the difference between each empirical value and the corresponding to the best fit, according to the theory) with the mobility reduction in each county. We estimate the mobility reduction for each county according to the data provided by *Google mobility* for the US –the only one of the four countries with municipal-level data available– on the percentage variation of time that people stay at home (see Materials and Methods section for data source and methodology).

In Fig. 4 we present the results as an histogram, where we can see a clear correlation between the reduced mobility and the the marginal deviation of *R*_0_. The higher the reduced mobility, the more negative the marginal deviation. In other words, the less the people move, the lower the value of *R*_0_ relative to the expected one for a given city size.

**Fig. 4.**
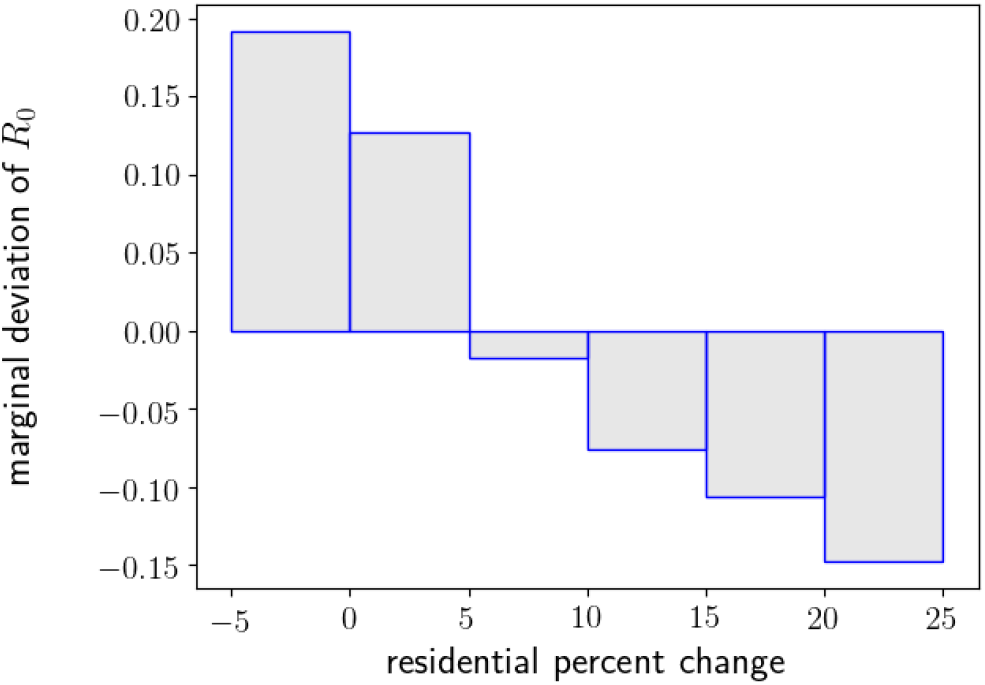
Average marginal deviation of *R*_0_ *vs* average mobility reduction for counties of the United States.

More generally, the marginal deviations may evidence the differences in social and political aspects regarding the COVID-19 problem in different countries. In any case, our results are a warning for large cities to act promptly, applying policies of social distancing and orders to stay home.

### *R*_0_ and population size

We have found that the size driven hypothesis, besides explaining many scaling demographic characteristic of urban centers (6), it also makes sense of the observed data regarding the COVID-19 epidemics. Moreover, after the fit of Eq. 5 for the four countries (Fig. 3), we obtain a universal scaling relation for the COVID-19 basic reproductive number ^2^

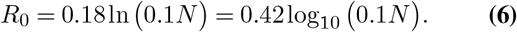

This simple logarithmic relation between *R*_0_ and the city size condenses both the theory and the observations regarding the spread of the COVID-19 pandemic in urban centers. Despite being simple, several conclusions can be made from it. It states that the mass action hypothesis –Anderson and May (10) call it proportionate mixing– is not generally valid, so the pattern of transmission depends on the city size. Moreover, Eq. 6 tell us quantitatively how its population size would impact a city in case of an epidemic. Indeed, it indicates that a multiplicative increase in population size results in the additive increase in the basic reproductive number. Therefore, the mass action hypothesis (*R*_0_ independent of *N*) overestimates the epidemic impact in small cities, while it underestimates for larger ones.

With respect to the non pharmacological measurements to fight the epidemic, like the stay at home recommendation or lockouts, they are normally based on city-independent basic reproductive number –implicitly following the mass action hypothesis. According to our findings, that would make some political actions overdone for some counties and inefficient for others. A more accurate strategy, however, should consider the present contribution scaling trend. And the same applies to reopening strategies.

We remark that the values of *R*_0_ obtained from our analysis and displayed in Fig. 3, including obviously the best fit of Eq. 6, are all within the range of the reported values in previous works (11–17).

In the following subsections we present three important conclusions that follow from the city population scaling of *R*_0_.

### Time dependent scaling

At the early stages of the epidemic, during the exponential phase, the time evolution of number of cases 𝒞 obeys (see Materials and Methods section for a complete theoretical analysis):

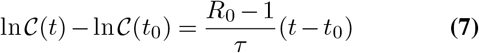

which, using Eq. 6 for *R*_0_ and *τ* = 8 days (18, 19), results in:

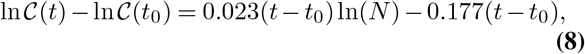

That is a power-law relation for the number of cases in term of the population size: 𝒞(*t*) *∝ N* ^*α*^, where the scaling factor *α* (*α* = 0.023*t* from Eq. 8) increases with time *t*. Figure 5 confirms this scaling for the US counties, while a recent empirical study shows a similar behavior for Brazilian cities (20). This is another evidence of the mass action hypothesis failure, since within it, *α* = 0 would be expected. In contrast, the exponent that increases over time provides further evidence in favor of the model, diverging from the mass action predictions as time increases.

**Fig. 5.**
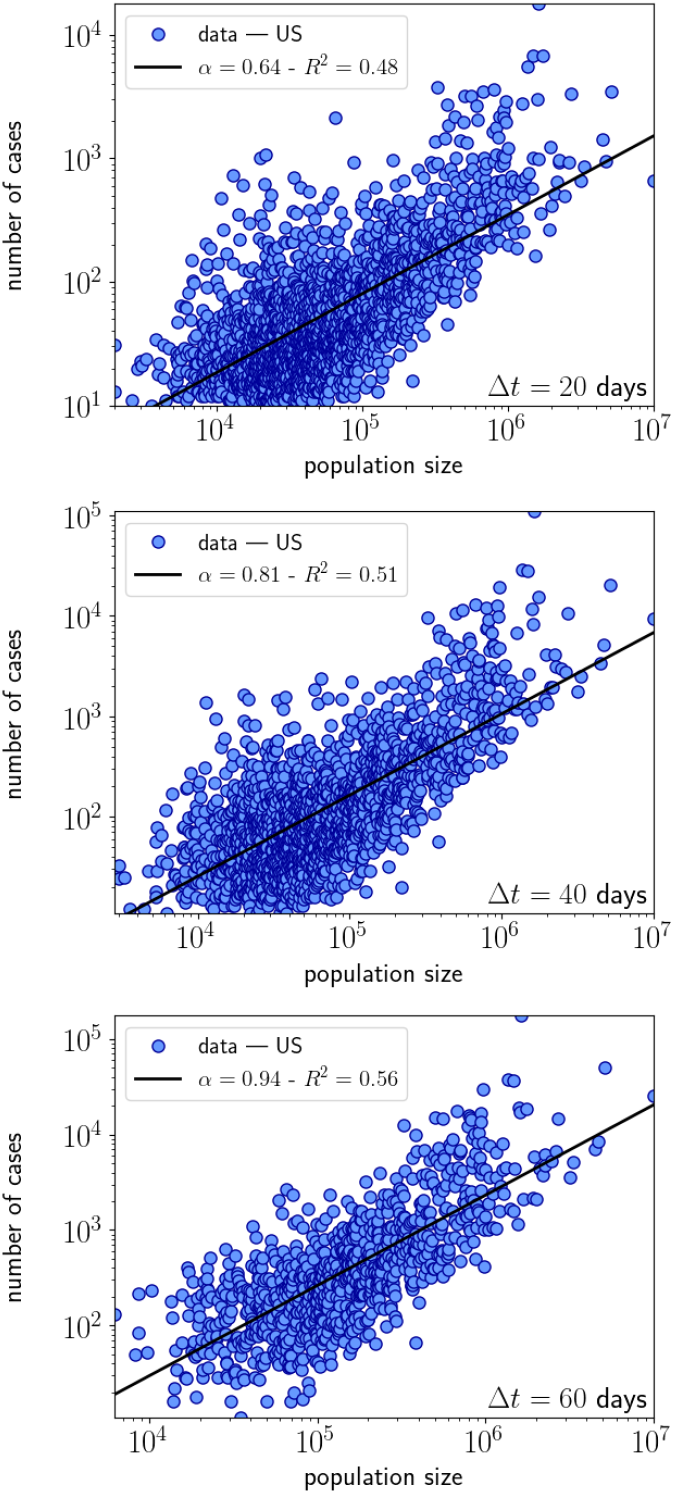
Relation between the number of confirmed cases and the population size of the US counties, at three different Δ*t* days (20, 40, and 60) after the 10th confirmed case.

### Epidemic peak

The so-called *peak* of an epidemic denotes the time and the value at that time when the number of active cases, *I*_*p*_, is at its maximum. Since a fraction of it requires hospitalization, a good regional/national health resources management should consider the way that number varies with the city size. According to our findings, the planning for a proportional distribution of ICU beds would lead to an underestimation of the necessary resources, with the danger of overloading the city’s health system.

The fraction of active cases at the epidemic peak, *I*_*p*_*/N*, is related to the basic reproductive number through the relation in Eq. 11 which, using Eq. 6, can be related to the city size, as we display in Fig. 6(a), for the theoretical expected fraction of active cases at the peak as a function of population size. Evidently, larger cities have relatively more active cases. More precisely, the *per capita* active cases grow approximately – fitting the exact expression– with ln(0.89*N*). It is worth mentioning that the theoretically predicted epidemic peak is hardly reached, because the non-pharmacological measures usually succeed in lowering the effective *R*_0_ before that.

**Fig. 6.**
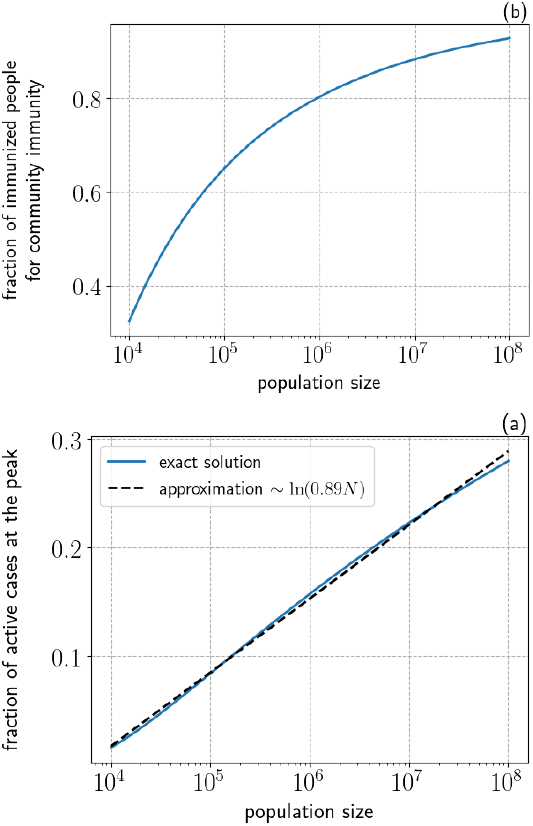
Theoretical expected fraction of (a) active cases at the epidemic peak and (b) people necessary for community immunity as a function of city population size.

### Community immunity

When a certain fraction of the population is immune –*i*.*e*. recovered from the infection–, the epidemic can not continue to evolve. At that point, any further outbreak would fade away. That condition is called *community immunization* and it is expressed mathematically in Eq. 12. Once more, by means of Eq. 6, we can express the fraction of people for community immunization, 𝒞^∗^*/N*, in terms of the city size, as shown in Fig. 6(b).

As the necessary fraction of the population increases fast with *R*_0_ and this in turn increases with size, it ends up being another epidemic quantity which scales with population size – in a rather more complex relation, as can be seen in Fig. 6. In a nutshell, larger cities would require more people immunized to achieve the desired *community immunization*. This not only affects the easing of restrictions eventually made based on sample draw of antibody prevalence, but it would have to be taken into account, once a vaccine is available. The economic downside of this aspect would be eventually counterbalanced by the city income, which scales with the population too (6).

## Materials and Methods

### Demographics

With municipal-level data of population density and population size for the United Sates (21, 22), Brazil (23, 24), Germany (25), and Portugal (26, 27), we fit a power-law function *ρ ∝ N* ^*λ*^ using a linear regression of the log-transformed variables, in the form ln(*ρ*) = *k*_1_ + *λ* ln(*N*), where *k*_1_ is a constant. We compare the result against the linear function *ρ ∝ N*, ln(*ρ*) = *k*_2_ + ln(*N*), and the constant function *ρ ∝*1, ln(*ρ*) = *k*_3_, where *k*_2_ and *k*_3_ are constants. We quantify the goodness of fit by computing the *R*^2^ for these three models, using log-transformed variables.

### Epidemic Model

To conduct our proposed analysis, which is to check how the COVID-19 transmissibility scale with the density/size of the urban population, we use the standard SIR model at the municipal level. We consider a population of size *N* divided in the following epidemiological compartments: susceptible *S*, infected *I*, and removed ℛ (recovered, immunized, or dead individuals). The dynamics of these compartments is driven by the following system of differential equations (4):

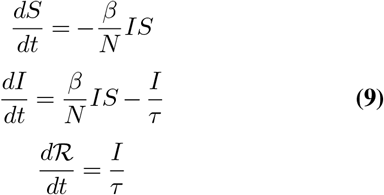

where *β* = *pC* is the transmission rate and *τ* is the infectious period. The *N* factor in the denominator makes *β* a disease only parameter, supposedly independent of the size or any other characteristic of the population. Called *mass action hypothesis* (4), *it assumes that the per capita* number of contacts is independent of the population size, resulting in similar patterns of transmission, whether it is a town or a large city (10). Now, defining the cumulative number of cases 𝒞 ≡ *N* − *S*, from Eq. 9 we got

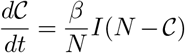

and

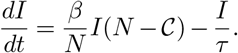

From the two above equations we have that

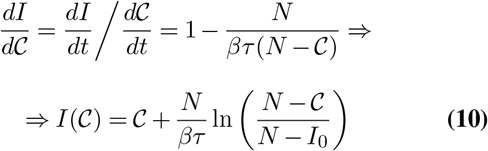

And from there, we get for the time derivative of 𝒞

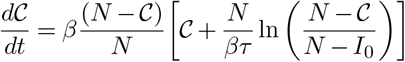

with the initial condition *I*(*t* = 0) = 𝒞(*t* = 0) = *I*_0_. At the early stages of the epidemic, when *I*_0_ *<* 𝒞 ≪ *N*, we can use a first-order approximation for the expression above to get

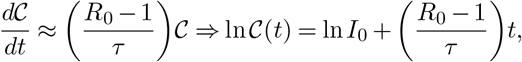

where *R*_0_ = *βτ* is the *basic reproductive number*. We use it here to estimate the value of *R*_0_ for each municipality as

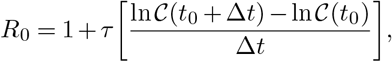

where *t*_0_ is the first day since the 10^*th*^ confirmed case, Δ*t* = 30 days to avoid any possible within-month measurement bias, and the infectious period is *τ* = 8 days (18, 19). For that purpose, we use the municipal level time series data of confirmed cases for the United States (28), Brazil (29), Germany (30), and Portugal (31). As for the mobility reduction for the US counties, we measure it during the epidemic in relation with the pre-pandemic standards. To do so, we estimate it with the average of *residential percent change from baseline* variable of *Google mobility reports* (32) over the priod [*t*_0_, *t*_0_ + Δ*t*].

#### Fit methodology

As explained in *Contact rate scaling theory* Section, we expect that *R*_0_ = *k*_1_ + *k*_2_ ln *x*, where *x* may be the population size or the population density (see *Demographics* for data source) and *k*_1_ and *k*_2_ are constants. We fit the relation using a linear regression with the log-transformed variable *x* and compare the result against the constant function *R*_0_ = *k*_3_, where *k*_3_ is a constant. We quantify the goodness of fit by computing the *R*^2^ for the two models.

#### Some predictions

The peak of an epidemic is when the number of active cases *I* is maximum. So, in that situation, we have

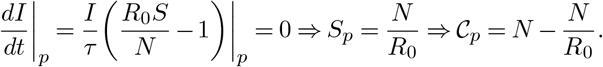

Inserting this result for 𝒞_*p*_ in the Eq. 10 for *I*(𝒞), the fraction of active cases during the peak is given by

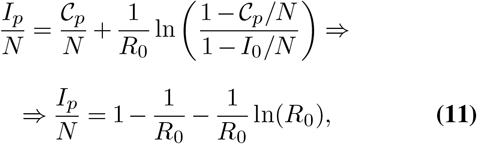

since *I*_0_ ≪ *N*.

In the equilibrium (community immunity), the cumulative number of cases is constant. So, the final fraction of infected persons is given the the solution of the transcendental equation

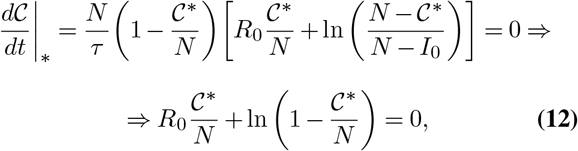

since *I*_0_ ≪ *N*.

## Data Availability

Data available from authors under request

## ACKNOWLEDGEMENTS

SG acknowledges CAPES for support, under fellowship #003/2019 - PROPG - PRINT/UFRGS, and URPP Social Networks of University of Zürich and Prof. Claudio Tessone for hospitality. S.G. also acknowledges Dr. Eduardo N. Warman for valuable comments.

## Supplementary Information Appendix

**Fig. S1.**
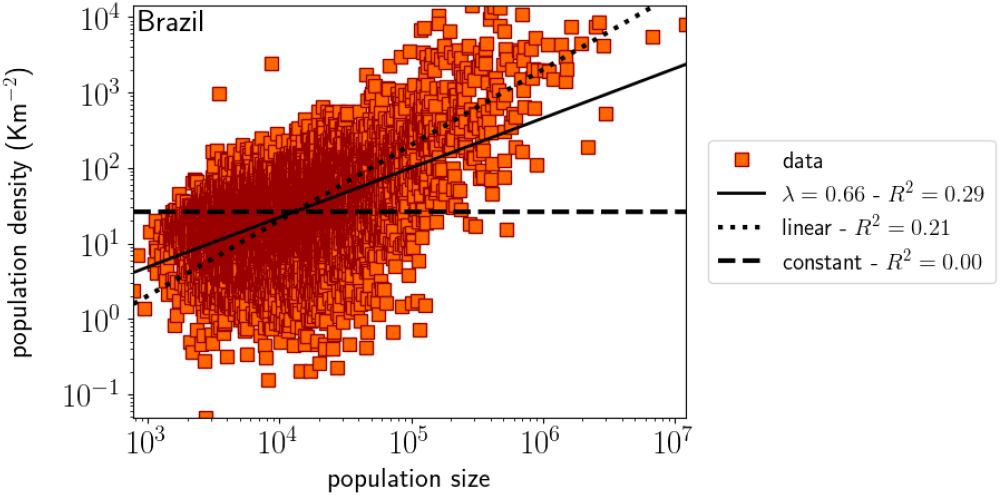
Population density (*ρ*) *vs* population size (*N*) at the municipal level for Brazil, represented by red squares (data). The solid line is a fit to a power law, *ρ∝N* ^*λ*^, while the dotted line is a fit to a linear relation, *ρ* = *kN*, with *k* = 0.002 Km^−2^. The dashed line corresponds to the average density. See Supplementary Materials for data source and fitting methodology.

**Fig. S2.**
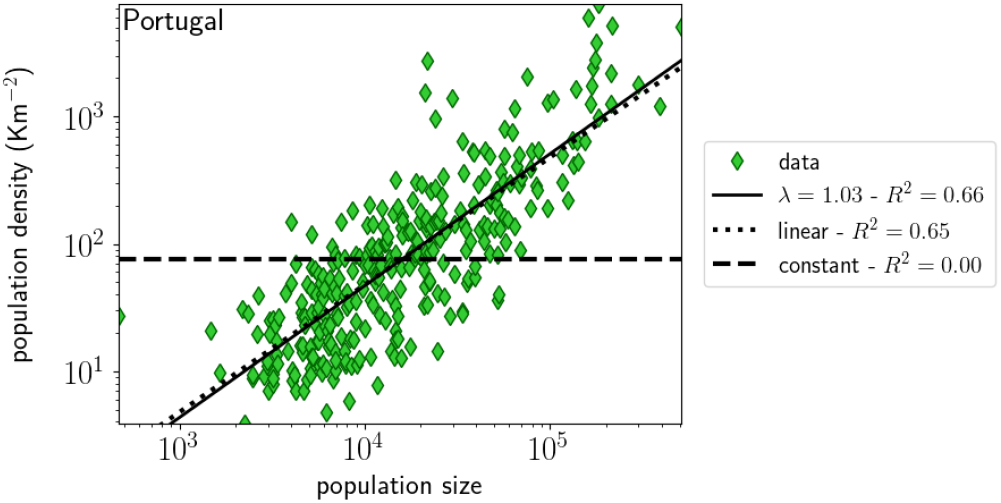
Population density (*ρ*) *vs* population size (*N*) for Portuguese municipalities, represented by green diamonds (data). The solid line is a power law fit, *ρ* ∝*N* ^*λ*^, while the dotted line is a fit to the *ρ* = *kN* relation with *k* = 0.00047 km^−2^. Both fits have almost equal correlation coefficient, *R* = 0.81. The dashed line represents the average density.

**Fig. S3.**
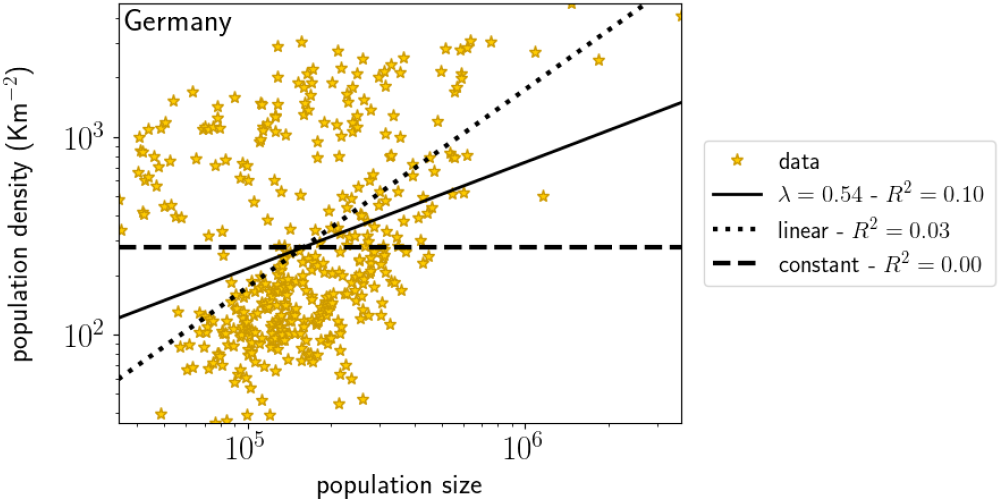
Population density (*ρ*) *vs* population size (*N*) at the municipal level for Germany, represented by yellow stars (data). The solid line is a power law fit, *ρ ∝N* ^*λ*^, while the dotted line are linear fit *ρ* = *kN*, with *k* = 0.0017 km^−2^. The dashed line corresponds to the average density

**Fig. S4.**
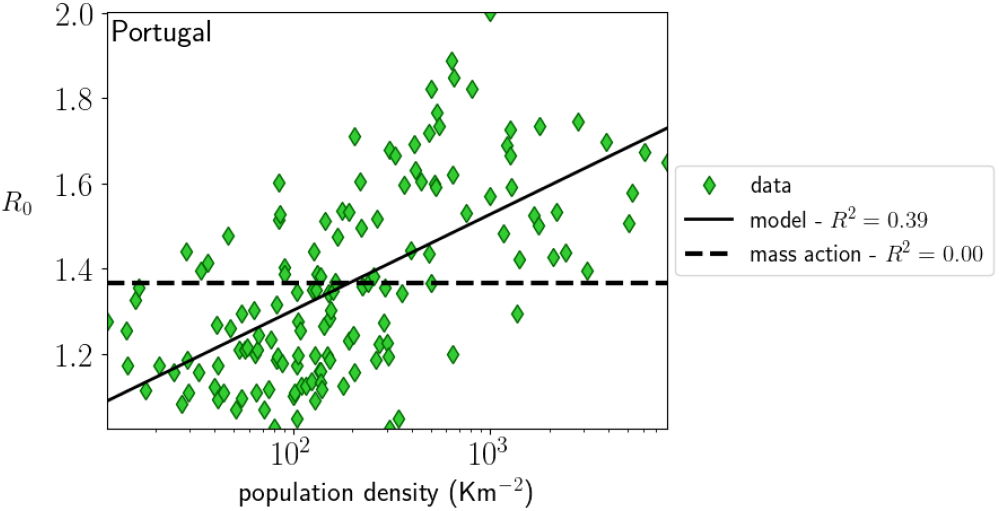
*R*_0_ *vs* the population density for different municipalities of Portugal.

**Fig. S5.**
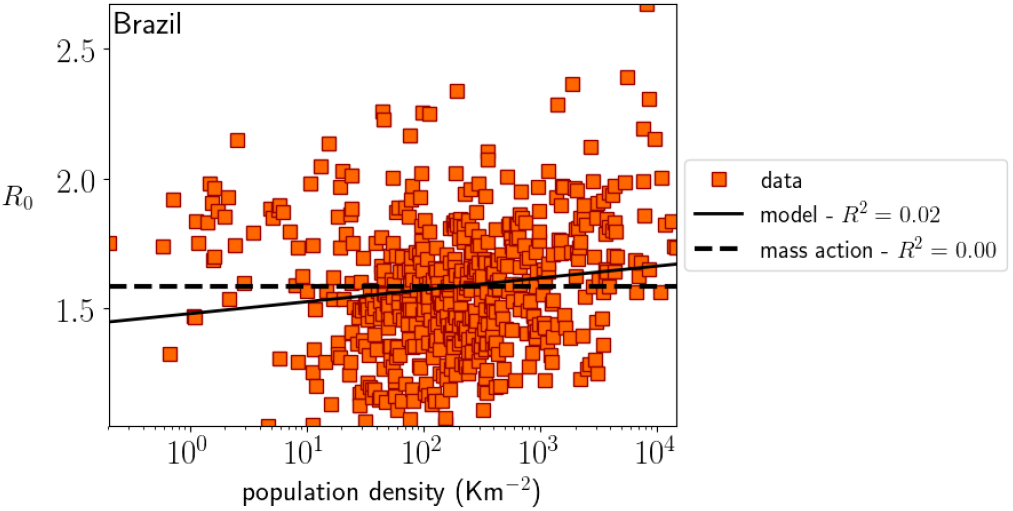
*R*_0_ *vs* the population density for different cities of Brazil.

**Fig. S6.**
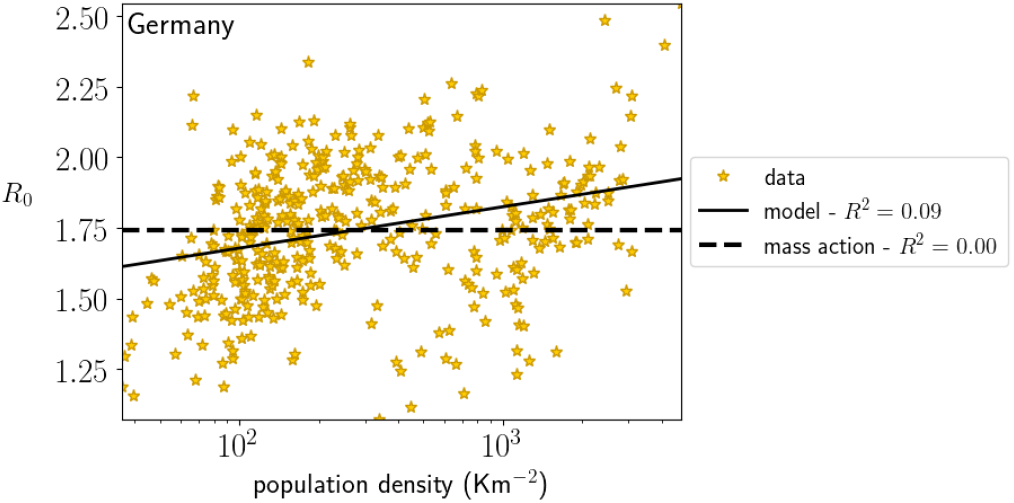
Comparison between *R*_0_ and the population density for different cities of Germany.

**Fig. S7.**
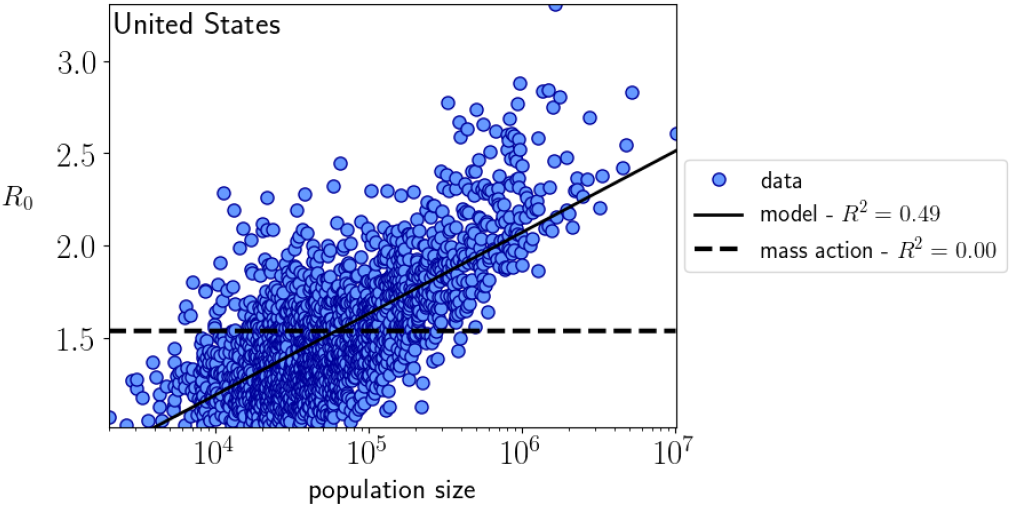
*R*_0_ *vs* the population size for different counties of the United States.

**Fig. S8.**
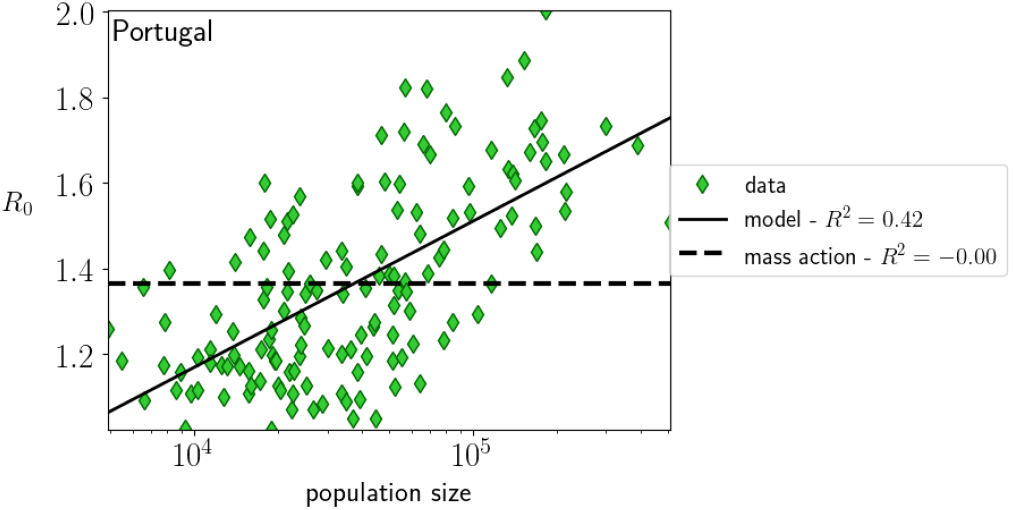
*R*_0_ *vs* the population size for different municipalities of Portugal.

**Fig. S9.**
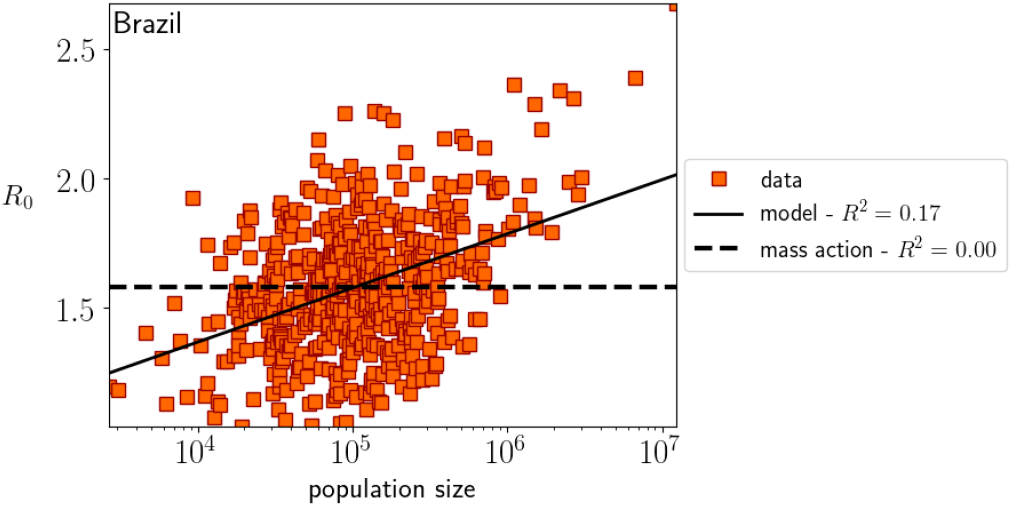
*R*_0_ *vs* the population size for different cities of Brazil.

**Fig. S10.**
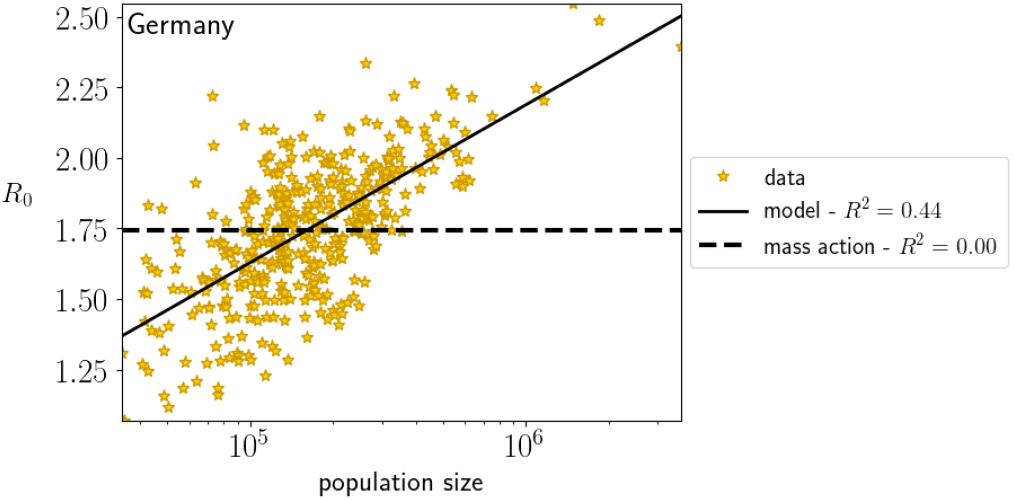
*R*_0_ *vs* the population size for different cities of Germany.

## Footnotes

1 It can be drastically attenuated or even suppressed by the use of masks, for example.

2 Our estimation method for *R*_0_ is valid for urban center bigger than *N* ≈ 3000, as it requires the number of cumulative cases to be small compared to the population size. For smaller towns, this is not always the case.

## Notes

### Competing Interest Statement

The authors have declared no competing interest.

### Author Declarations

The present research was conducted using COVID-19 and demographic data solely from public sites. No human was involved in this research.

